# Bayesian Nonparametrics for Normative Modelling in Multiple Sclerosis via Modularised Inference

**DOI:** 10.64898/2026.05.10.26352835

**Authors:** Bernd Taschler, Thomas E. Nichols, Habib Ganjgahi

## Abstract

Normative models produce per-subject deviation scores that feed directly into down-stream analyses, but typical pipelines (i) treat confounders with ad-hoc or purely linear adjustments, and (ii) pass point estimates of deviation scores directly to the down-stream model, ignoring uncertainty.

We propose an integrated, two-module Bayesian framework that aims to address both limitations. A normative module based on Bayesian Additive Regression Trees (BART) flexibly captures non-linear effects and higher-order interactions while marginalising over image-quality variables via counterfactual averaging. Crucially, we define individual deviation as 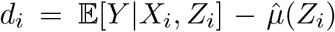 with 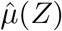 the feature-conditional population mean, not as a residual (*r*_*i*_ = *Ŷ*_*i*_ − *Y*_*i*_). A SoftBART survival model then ingests the full posterior distribution of deviation scores via a cut-posterior construction, propagating upstream uncertainty while blocking feedback from the outcome model.

Across challenging simulations and a large clinical data set of multiple sclerosis patients (N*>* 8k), the integrated approach yields better calibration, prediction accuracy and time-varying hazard separation between groups than a two-step plug-in Cox regression model. Modularised inference with BART-based normative deviations improves both flexibility and uncertainty quantification, and extends naturally to other outcomes beyond survival.

## 1 Introduction

Over the past decade, neuroimaging has increasingly shifted from explaining average group differences to understanding heterogeneity at the level of the individual. Conventional case–control analyses are poorly suited to this goal as they characterise mean effects in diagnostic groups, but provide little guidance on where a given subject lies relative to an appropriate reference population and how atypical their brain phenotype is. In contrast, normative modelling explicitly targets subject-level variability by learning population reference distributions and quantifying individual deviations from these norms, thereby aligning with broader aims in psychiatry and neurology to move beyond group averages and towards personalised inference (Wolfers et al., 2018; Bethlehem et al., 2022; Rutherford et al., 2022; Fraza et al., 2024, 2025).

### Background and motivation

Normative modelling has become an influential statistical framework for characterising individual variability by comparing observations against a population-level reference. By estimating population-level distributions of imaging-derived phenotypes conditional on relevant covariates—such as age, sex, disease history, etc.—normative models provide subject-specific reference metrics that quantify deviation from expected biological patterns.

In the literature, “normative modelling” has been used to refer to three different areas of research: brain age estimation, lifespan modelling and understanding population heterogeneity. Brain age studies are focused on predicting chronological age from imaging features and other covariates. The residual difference between predicted and observed age (the brain age gap) is used as a scalar summary indicator for downstream analyses. Early approaches used linear regression to model brain morphometric features as functions of age and sex (Franke et al., 2010; Liem et al., 2017; Cole et al., 2017), while advanced machine learning and deep learning models have recently become increasingly popular (Franke and Gaser, 2019; More et al., 2023).

Lifespan modelling differs from brain age estimation in that its goal is to estimate population reference trajectories of brain phenotypes across age, and provide centile scores analogous to anthropometric growth charts. Flexible statistical models such as generalised additive models (GAMs; (Dinga et al., 2021)), possibly including modelling of conditional distribution skew and kurtosis (GAMLSS), have been proposed to improve centile estimation and site harmonisation. The landmark study by Bethlehem et al. (2022) used GAMLSS to model reference trajectories from infancy to old age. Another study has used normative models to generate lifespan-wide normative maps of cortical thickness and other structural phenotypes (Rutherford et al., 2022). These charts provide a way to situate an individual vs a population baseline, but are not designed for individual prediction of disease-related outcomes.

Into the third category fall models that map individual deviations from a population norm while explicitly modelling multiple sources of variation (biological and nuisance) to capture population heterogeneity and enable subject-level inference. Early work proposed Gaussian process (GP) regression as a non-parametric Bayesian modelling approach that naturally provides uncertainty estimates (Marquand et al., 2015), and was further extended to multi-task GPs for joint modelling of multiple imaging modalities (Kia and Marquand, 2018). Hierarchical Bayesian regression has been applied in multi-site studies to pool data across cohorts while accounting for site effects (Kia et al., 2020; de Boer et al., 2024). Other studies have used normative models to characterise atypical brain structure and function (Wolfers et al., 2018; Fraza et al., 2024, 2025). These models provide subject-level deviation scores with uncertainty, disentanglement of variance sources, and flexibility to integrate into downstream inference (e.g., classification, clustering, survival models).

A systematic comparison of modelling strategies by Bozek et al. (2023) found that sample size, model choice, and evaluation criteria all substantially affect the accuracy of derived percentiles, especially in the distribution tails. Most normative modelling pipelines rely on residual-based deviation scores, which are sensitive to noisy observations and confounded by nuisance variables such as image quality. Moreover, few approaches include all available covariates in the model and fully account for potential confounding.

Finally, downstream analyses such as disease prediction, classification or survival modelling, are commonly conducted in a two-step fashion, ignoring the uncertainty associated with deviation scores, which might lead to spurious findings, inflated false positives and overconfident estimates. Bayesian models estimate the normative mapping and downstream parameters together, so that uncertainty is integrated in a single posterior (Kia et al., 2020). However, model complexity and computational burden make fully joint models very challenging and in many scenarios practically infeasible. Instead, approximate Bayesian and variational approaches can be used to return a posterior approximation or multiple samples and thus provide uncertainty estimates that can be propagated into downstream models (Fraza et al., 2021).

### Our contribution

In this work, we build on these advances and introduce a hierarchical Bayesian framework for normative modelling with modularised inference. Our method has the following key features: (i) Cut-posterior inference: We implement a modularised inference scheme to propagate uncertainty from the normative module into a downstream outcome model (e.g., survival analysis), while explicitly blocking feedback to preserve robustness when the second module is potentially misspecified. (ii) Subject-level deviation scores: We define individual normative deviation scores as difference between predictions and the estimated population mean conditional on covariates of interest (e.g., disease severity), not as residuals from observed data. (iii) Normative modelling: We use Bayesian Additive Regression Trees (BART) to model conditional distributions of imaging phenotypes together with all available covariates, capturing non-linear relationships, interactions, and potential confounding via image quality and scanner effects. To our knowledge, this is the first use of BART and modularised inference in a normative modelling context.

This modular architecture maintains a coherent probabilistic interpretation without requiring full joint inference, and it generalises beyond survival outcomes to support classification, regression, or clustering tasks.

### Outline

The remainder of the paper is organised as follows: In Section 2, we introduce the hierarchical Bayesian framework, detailing the normative module based on BART, our expectation-based definition of deviation scores, and the modular integration with a downstream survival model using cut-posterior inference. Section 3 presents empirical results on a large MS dataset, as well as controlled simulations designed to evaluate model performance in comparison to a traditional plug-in survival model. We conclude in Section 4 with a summary of key findings, implications of our modelling choices, and potential extensions.

## 2 Methods

The following subsections introduce the modular inference framework and define the normative and survival models, respectively. At a glance, key modelling concepts are summarised graphically in Figure 2.

### 2.1 Modularised Inference via the Cut-Posterior

In traditional Bayesian models, all parameters and latent variables are estimated with a single unified posterior distribution. The ideal scenario when using normative methods together with downstream models would thus involve fitting a single big model. However, with large neuroimaging datasets this is often infeasible in practice due to model complexity and scalability issues.

Large, complex models are often composed of distinct submodels. In this case joint inference enables the flow of information between submodels but assumes that all components are correctly specified and coherent within a joint generative framework. However, in practice, one component may be misspecified, overly simplistic, reliant on noisy or biased data, or computationally infeasible to integrate with a complex upstream model.

To address these challenges, we adopt a modularised inference strategy via the cut posterior (Plummer, 2015; Jacob et al., 2017; Chakraborty et al., 2022; Liu and Goudie, 2025). This approach decouples inference between model components while preserving uncertainty propagation from upstream to downstream modules. Specifically, we estimate posterior distributions for the parameters of the normative model (*θ*_1_) using data *D*_1_. Posterior samples of individual deviation scores *d*_*i*_ are then used as inputs to a downstream survival model with parameters *θ*_2_ and time-to-event data *D*_2_. Crucially, we cut feedback from the survival model to the normative model, i.e. the posterior of the normative model is treated as fixed, and its uncertainty is propagated forward, but not updated in light of the survival data.

Formally, this corresponds to making an assumption that the posterior factors as:

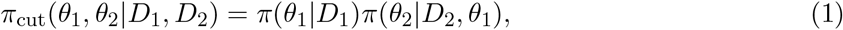

which is generally referred to as the cut-posterior distribution. This framework avoids updating *θ*_1_ based on *D*_2_, but accounts for uncertainty in *θ*_1_ when estimating *θ*_2_.

This is particularly desirable when feedback from the downstream model is either negligible (thus improving computational efficiency) or undesirable (e.g., due to misspecification). The trade-off is a departure from full Bayesian coherence—since the cut posterior no longer corresponds to a single joint probability model—but in exchange for increased robustness and interpretability in sequential modelling tasks. A schematic comparison of full joint versus modularised inference is shown in Figure 1.

**Figure 1:**
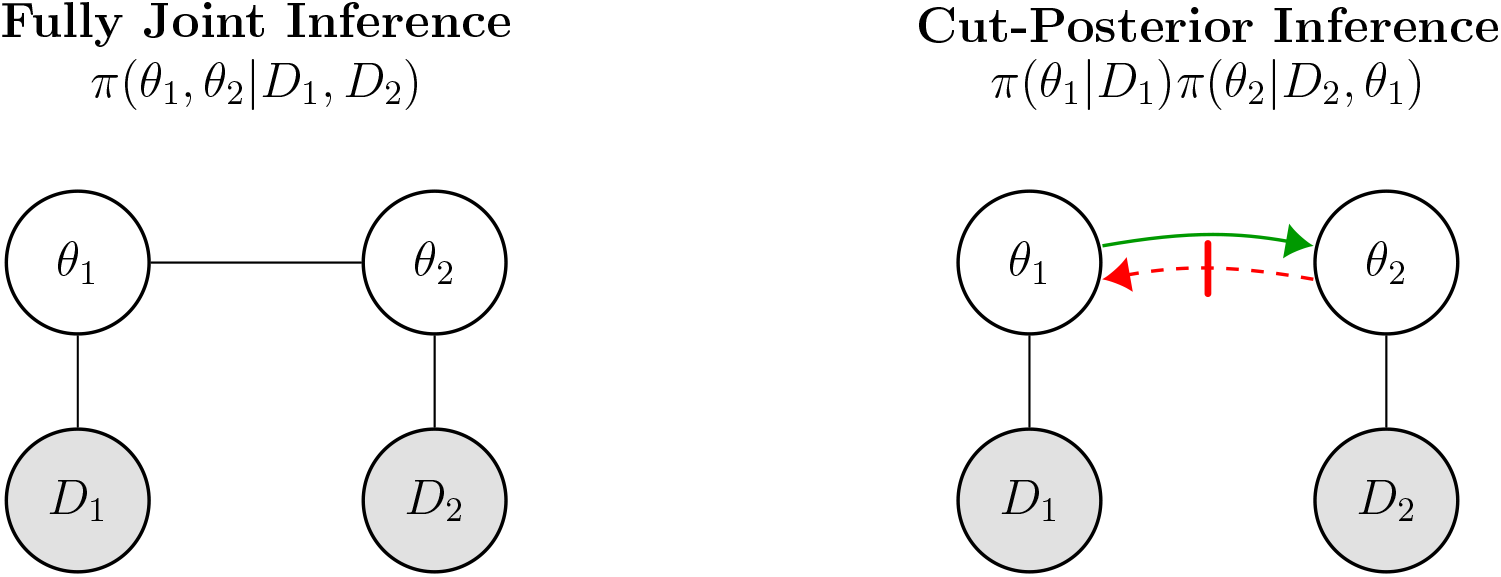
Schematic comparison of fully joint inference (left) and cut-posterior (modularised) inference (right). Solid arrows indicate active information flow; the red dashed arrow indicates feedback that is blocked in the modular approach. Filled nodes represent observed data, hollow nodes are model parameters.

### 2.2 Normative Model (Module 1)

We model normative variation via Bayesian Additive Regression Trees (BART). This flexible, non-parametric Bayesian ensemble method allows for a robust approximation of complex, potentially non-linear relationships and interactions without overfitting (Chipman et al., 2010; Hill et al., 2020). Unlike classical regression trees, BART produces posterior distributions over predicted outcomes, facilitating calibrated uncertainty quantification.

Let *y*_*i*_ denote the quantity of interest for subject *i*, for example, an imaging-derived phenotype (IDP) such as normalised brain volume (NBV). Further, let 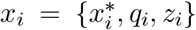 be the vector of covariates, including demographic and clinical covariates 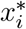, potential confounders and nuisance variables *q*_*i*_, and key feature variables *z*_*i*_. Any discrete (or discretised) covariate of interest could be considered as feature, on which the normative outcome is then conditioned on (see definition of deviation scores below). The standard BART model is defined as

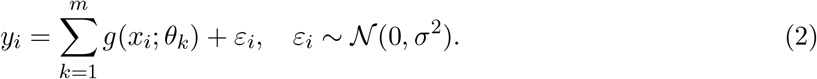

Here, each *g*(·; *θ*_*k*_) is a shallow regression tree with parameters *θ*_*k*_ = *{T*_*k*_, *M*_*k*_, *ψ}*, including structure parameters *T*_*k*_, terminal node (leaf) parameters *M*_*k*_ and priors *ψ*. For simplicity, we denote the set of BART-specific parameters collectively as Θ. Trees are constrained through regularization priors to have a small effect on the overall prediction. Specifically, BART places a prior over tree depth (penalising complex trees), a prior over splitting variables and rules, and a shrinkage prior (typically Gaussian) over the leaf values. The sum over *m* trees defines a rich ensemble prior. The model parameters are estimated using a Bayesian backfitting Markov Chain Monte Carlo (MCMC) algorithm that iteratively updates each tree while holding the others constant (Chipman et al., 2010).

#### Adjustment for confounders via counterfactual marginalization

A probabilistic model such as BART defines a posterior predictive distribution over outcomes *Y*, conditional on input covariates *X*. Such models naturally allow for sampling under arbitrary covariate settings, including those not observed in the training data—provided that the observed data sufficiently cover the entire covariate space to avoid extrapolation.

Consider the effect of an image quality measure (IQM), for example, the contrast to noise ratio (CNR). In classical linear regression, predictions of the outcome would be made conditional on a default value, e.g. the average CNR^1^. With BART, for each subject, we can predict the outcome under a range of (discretised) CNR values, average over those to obtain the posterior estimate and then over subjects to compute the expected population mean. See panel A in Figure 2 for a schematic comparison.

**Figure 2:**
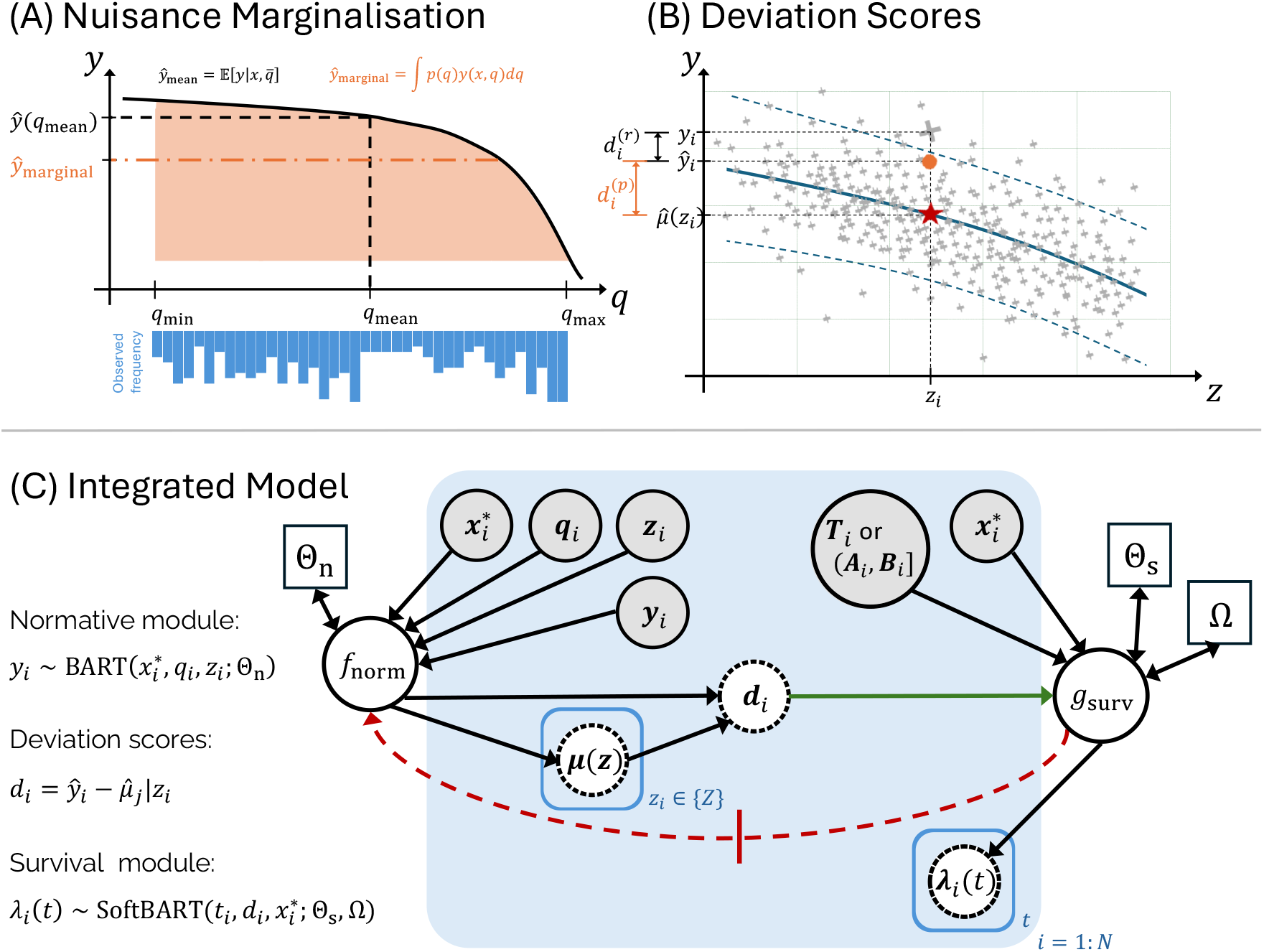
Schematic representations of key modelling concepts. (A) Comparison of expectations based on a population-averaged nuisance variable *q* and marginalisation over *q*. (B) Comparison of residual-based (*d*^(*r*)^) and prediction-based (*d*^(*p*)^) deviation scores. (C) Overview of the modularised inference approach, propagating uncertainty (green arrow) from the normative module on the left to the survival module on the right, while cutting feedback in the reverse direction (red dashed arrow). Circles represent random variables, shaded variables are observed, squares represent model parameters, dashed nodes are estimated.

Based on the posterior distribution, we can predict new data to obtain a posterior predictive distribution *p*(*Y* |*X*^∗^, *q*). For any value of the nuisance variable *q* ⊂ *X* one can thus compute counterfactual predictions by varying *q* while holding the remaining covariates *X*^∗^ = *X* | *{q}* fixed. Assuming further that *q* is discrete (or discretised), we can marginalise over nuisance variables via the sum over posterior draws:

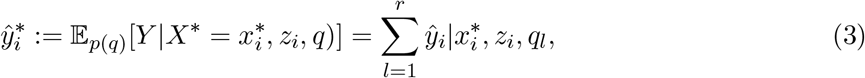

where *p*(*q*) is the empirical distribution of the nuisance variable, discretised into *r* bins. In real data applications (see Section 2.5), it may be necessary to marginalise over multiple nuisance variables *q* = *{q*_1_, …, *q*_*s*_*}*. In this case, *p*(*q*) can be approximated by creating an *s*-dimensional grid over quantiles of each nuisance variable, computing the counterfactual outcome for each grid point, weighted by the observed frequency of each (discretised) combination of nuisance values in the data. This marginalisation produces predictive expectations that are robust to nuisance variation and reflects integration over uncertainty in *q*. It is not a preprocessing step but a part of the posterior computation when fitting the model.

#### Deviation score definition

We define each subject’s deviation scores *d*_*i*_ as a de-noised individual subject prediction relative to the expected population mean conditional on a key covariate of interest^2^, for example, a measure of disease severity such as the Expanded Disability Status Scale (EDSS) in MS. Specifically,

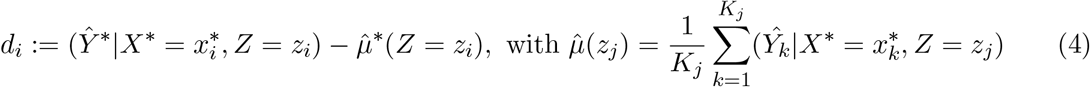

where 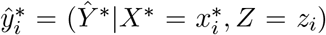 is the posterior mean of the outcome (e.g., NBV) for subject *I* from Eq. (3), and 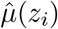 is the expected population mean over all subjects *K*_*j*_ at the same level of the feature covariate *Z* (e.g., EDSS). Panel B in Figure 2 shows a graphical comparison between these two definitions of deviation scores.

This formulation contrasts with residual-based scores which traditionally define deviation as *ŷ*_*i*_ − *y*_*i*_; for example, the brain age gap or brain volume cut-offs as used in Sormani et al. (2017).

In summary, our formulation ensures (i) marginalisation over nuisance variation, (ii) conditioning on key feature covariates, (iii) no mixing of point estimates with noisy data, and (iv) principled uncertainty propagation.

### 2.3 Survival Model (Module 2)

Our Bayesian survival model with interval-censored events and a BART covariate model is defined as follows. Let *x*_*i*_ ∈ ℝ^*p*^ denote the vector of observed covariates for subject *i, i* = 1, …, *N*. The true event time *T*_*i*_ ∈ (0, ∞) is assumed to lie within an interval (*A*_*i*_, *B*_*i*_], where *A*_*i*_ and *B*_*i*_ are scheduled clinical assessments, for example. In the case of exact (uncensored) event times, we have *A*_*i*_ = *B*_*i*_ = *T*_*i*_.

Let *g*(*t, x*_*i*_) denote a nonparametric time- and covariate-dependent predictor function, which is modelled using SoftBART (Linero, 2022)—a smooth generalisation of the standard BART model where tree-based splits use soft logistic transitions rather than hard thresholds.

We model the hazard function *λ*_*i*_(*t*) as

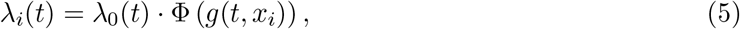

where *λ*_0_(*t*) is a parametric baseline hazard function and Φ(·) is the standard normal CDF. When fitting the model we assume a constant baseline hazard, i.e. *λ*_0_ = Ω, the rate of an exponential distribution.

The corresponding survival function is

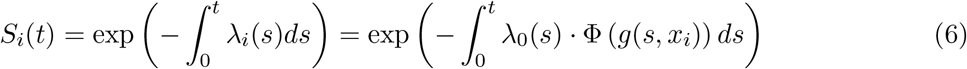

The likelihood contribution of subject *i*, whose event is known to lie within the interval (*A*_*i*_, *B*_*i*_] is given by Pr(*T*_*i*_ ∈ (*A*_*i*_, *B*_*i*_]) = *S*_*i*_(*A*_*i*_) − *S*_*i*_(*B*_*i*_). Assuming independence, the full likelihood is the product over all *N* subjects, i.e. 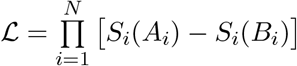.

#### Data augmentation for efficient sampling

Our implementation of the SoftBART survival model follows closely the approach proposed by Basak et al. (2022). Specifically, we adopt a two-layer data augmentation scheme to enable evaluation of the integral in the survival function.

The first layer involves sampling latent event times from a thinned non-homogeneous Poisson process (NHPP). Let 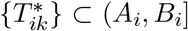 denote the set of proposed latent event times for subject *i*, indexed by *k* = 1, …, *K*_*i*_, and obtained by sampling from a standard homogeneous Poisson process with baseline intensity *λ*_0_(*t*) and thinning each point with the acceptance probability Φ (*g*(*t, x*_*i*_)). The sampled event time is 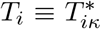 with *κ* indicating the index of the first accepted point in the NHPP and 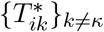 correspond to rejected points. This approach accounts for interval-censoring and allows for simpler, slice-sampling based updates of the model parameters.

The second augmentation layer uses latent continuous responses to enable sampling from the intractable integral in Equation (6). The Bayesian back-fitting algorithm of Chipman et al. (2010) for BART requires the response function *g* in the form of a semi-parametric Gaussian regression (*Y* = *g*(*t*, **x**) + *ε*). However, *g* in Equation (5) appears inside the probit terms Φ(*g*(*T*_*i*_, **x**_*i*_)). Let *Z*_*ik*_ ∈ ℝ denote latent Gaussian variables, corresponding to each time point from the first augmentation scheme. These variables are sampled from truncated normal distributions with unit variance and serve as pseudo-responses for the SoftBART backfitting algorithm of Linero et al. (2022).

Basak et al. (2022) show that, with these two layers of data augmentation, the probit terms Φ(·) in the likelihood cancel out, resulting in the joint likelihood

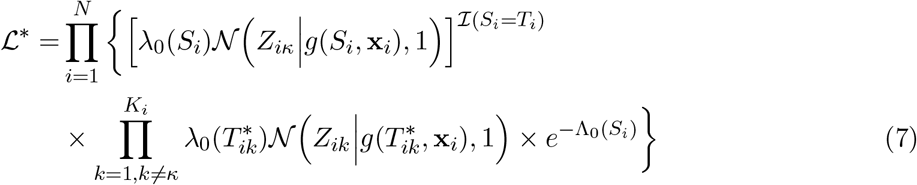

where *S*_*i*_ = *T*_*i*_ if the survival time is observed and *S*_*i*_ = *A*_*i*_ in the right-censored case; 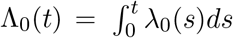 denotes the baseline cumulative hazard function of the survival times.

We assume a constant baseline hazard *λ*_0_(*t*) = Ω and place a Gamma prior with shape and rate parameters estimated from the data. For an in-depth discussion of the choice of baseline hazard, see Linero et al. (2022).

The fully augmented model consists of the following components:

1. Hyperparameters: BART parameters Ψ ∼ *p*(Ψ) and baseline hazard parameter Ω ∼ Gamma(*α, β*)
2. Predictor function: *g* ∼ SoftBART(Ψ)
3. Event time model: 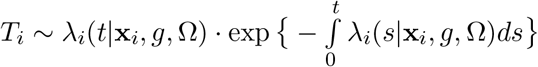
4. Latent event times: 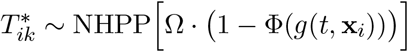
5. Latent responses: 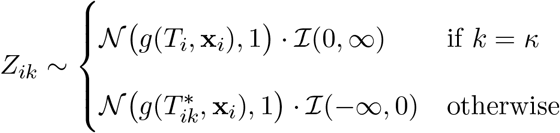

#### Model fitting and sampling

Posterior updates of the baseline hazard are computed via closed-form expressions using the augmented data. We initialise the baseline hazard 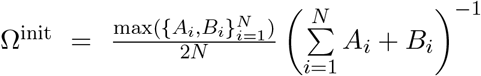and update the baseline hazard via Ω ∼ Γ(*α* + *N* + Σ_*i*_ *K*_*i*_, *β* + 2 Σ_*i*_ *T*_*i*_) with hyperparameter priors *α* = 1 for shape and *β* = 1*/*Ω^init^.

For the SoftBART tree model, we fit a total of *m* = 50 trees and keep all other parameters at their default values (Linero, 2022). Pseudo-code for the MCMC routine is presented in Algorithm 1.

Note that if *T*_*i*_ is observed (uncensored) then *A*_*i*_ = *B*_*i*_ and the while-loop is not evaluated; similarly, the second for-loop then is only evaluated for *l* = 1, …, *K*_*i*_.

#### Integration of posterior deviation scores into the survival model

Within the cut-posterior framework, each subject’s covariate vector *x*_*i*_ is extended by appending the deviation score *d*_*i*_. This score is updated at every MCMC iteration with draws from the posterior distribution of the normative model. The process is as follows: After fitting the normative module, we store *R* posterior draws 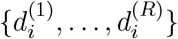 for each individual *i*. During fitting of the survival model, at each iteration *r*, we update the covariate vector as

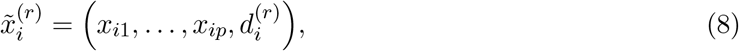

where *x*_*i*1_, …, *x*_*ip*_ are the observed clinical and demographic covariates. The survival model’s regression surface 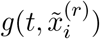 is then evaluated using this extended covariate vector when computing the likelihood and performing the back-fitting updates for the SoftBART components. This procedure ensures that uncertainty in *d*_*i*_ across posterior draws is propagated into the hazard function estimates, rather than fixing *d*_*i*_ to a single point estimate. By construction, the MCMC updates for the normative model are not revisited during survival model fitting, preserving the modularization and avoiding feedback from the survival likelihood as described in Section 2.1.

##### Algorithm 1

Sampling from the augmented model.

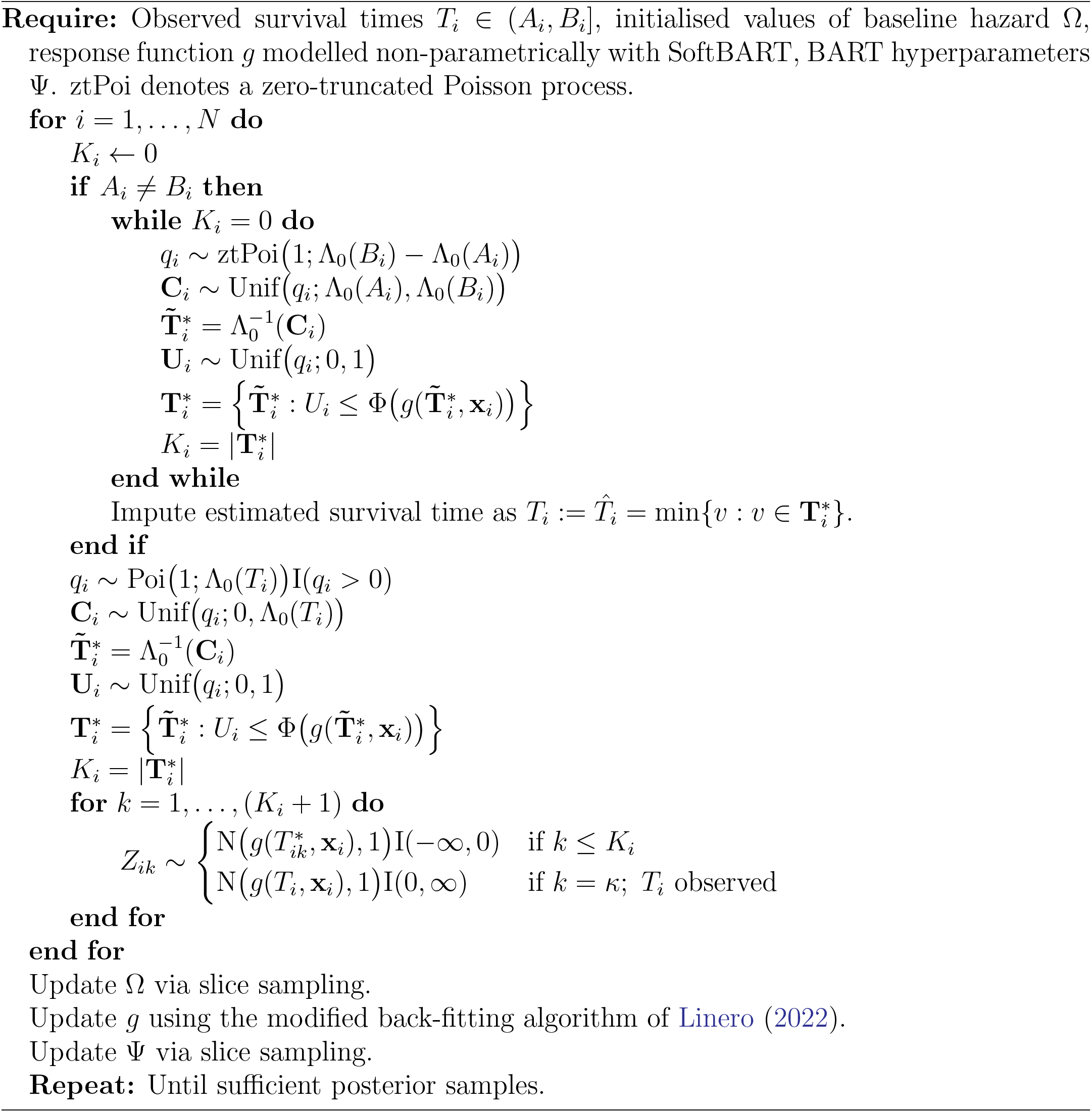

#### Integrated model overview

Figure 2 (panel C) represents a schematic representation of the full model.

### 2.4 Simulations

To evaluate the proposed pipeline under controlled conditions, we designed a two-stage simulation framework that reflects the structure of the normative and survival modules.

This simulation design deliberately embeds the survival signal in a latent, noise-free phenotype (*Y*) that is only observed indirectly and noisily 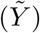 through the normative model. This mirrors the intended real-world application, where imaging-derived measures are affected by nuisance variation (e.g., site effects, measurement error, etc.). The survival risk depends on the latent signal, so accurate uncertainty propagation from the normative model into the survival model is critical for well-calibrated risk estimation. The setup therefore directly tests whether the cut-posterior approach improves calibration and predictive performance over a standard two-step method that ignores deviation score uncertainty.

#### Normative model data generation

We generate *N × p* feature vectors *x*_*i*_ by sampling inde-pendently from a *p*-dimensional unit hypercube, with *p* = 5 and *i* = 1, …, *N*. The underlying noise-free response was defined using Friedman’s test function:

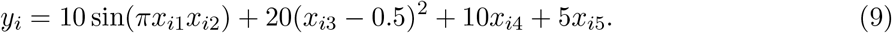

To emulate realistic noise and nuisance variability, we add to each observation four additional noise covariates, i.e. *x*_*i*6_ ∼ *U* (1), *x*_*i*7_ ∼ *N* (0, 1), 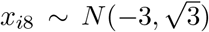, *x*_*i*9_ ∼ *Exp*(3), and define the noisy observed response as 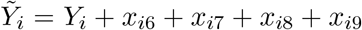. We generated independent training and test sets of size *N*_train_ = *N*_test_ = 1, 000.

#### Survival model data generation

To simulate survival times, we constructed individual-specific shape parameters *α*_*i*_ that depend on the informative covariates *{x*_*i*1_, …, *x*_*i*5_*}* and the latent noisefree phenotype *y*_*i*_:

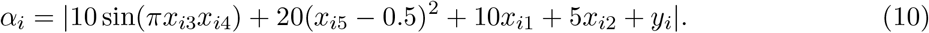

Survival times were then drawn from a Gamma distribution as *T*_*i*_ ∼ Γ(*α*_*i*_ + 0.1, *λ* = 6), ensuring that event risk is determined jointly by the covariates and the latent phenotype, while the model itself only observes the noisy outcome 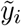. For simplicity, no right- or interval-censoring was applied in these experiments.

We create true group labels of ‘high’, ‘medium’ and ‘low’ (*g*_*i*_ ∈ *{*1, 0, −1*}*) using standardized *z*-scores of the noise-free outcome 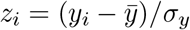,

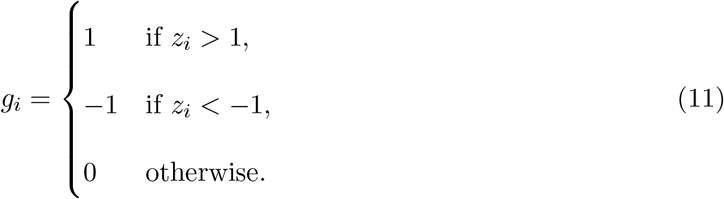

#### Model fitting

The normative model was fitted using BART with 200 trees, 10k burn-in, and 100k posterior draws with 1*/*10 thinning, resulting in 10k effective samples. The survival model was fitted using SoftBART with the same MCMC parameters but using the SoftBART default of 50 trees, and 50 evaluation time points over the unit interval. In the cut-posterior integration, deviation scores were sampled from the normative model’s posterior at each MCMC iteration of the survival model as described above.

#### Model comparison

For benchmarking, we also implemented a standard Cox proportional hazards (CPH) model using the same input features. In a two-step, plug-in approach, deviation scores were computed as their posterior mean from the normative model, without uncertainty propagation.

### 2.5 Real Data Application

We applied our proposed framework to a large cohort of individuals with multiple sclerosis (MS) from the NO.MS database (Dahlke et al., 2021), the largest aggregated resource of randomised clinical trial data in MS to date. It integrates 39 phase II and III clinical trials (2003–2021), including corresponding open-label extension phases. All contributing studies were approved by relevant ethics committees and conducted in accordance with the Declaration of Helsinki and Good Clinical Practice. Results from individual trials have been published elsewhere. All data have been de-identified using a risk-based anonymisation framework (Mallon et al., 2021; Delbarre et al., 2022). From the whole database, nine studies were selected based on availability of protocol-defined standardised clinical assessments and regular MRI acquisitions, resulting in a total sample size of *N* = 8, 324.

Few studies have applied normative methods in the context of MS. Early work by Sormani et al. (2017) categorised NBV based on residuals from a simple linear regression model. More recently, Garbarino et al. (2024) used a Gaussian Process Progression model (GPPM) to learn the typical disease trajectory and assess individual progression rates. The study used a traditional Cox proportional hazards model in post-hoc analysis to quantify risk differences between groups, ignoring uncertainty in normative deviation scores. While Cox proportional hazards regression is typically used in survival modelling, we developed a richer Bayesian time-to-event model. In most clinical settings, including MS, events are logged only at scheduled patient visits, and hence the event is only known to have occurred between two time points, often 3–6 months or more apart. Further, there are many covariates that could have complex effects on survival (e.g., non-linear relationships or interactions). Hence we implemented a semi-parametric Bayesian survival model that accommodates interval-censored time-to-event data with a BART covariate model that allows non-linear, time-varying effects.

We modelled individual normative deviations in total normalised brain volume (NBV) and used these deviation scores to predict risk of disability worsening in a downstream survival model. We demonstrate that our method yields better-calibrated and more interpretable risk profiles than conventional two-step approaches, both in real-world and simulated data settings.

Our modularised inference approach can be transferred to other settings by first obtaining a posterior distribution from the normative module and computing individual deviation scores, and then using the full posterior of deviation scores to formulate a downstream model (including survival, clustering, classification, prediction or other tasks).

#### Normative model outcome and covariates

For the normative module, normalised brain volume (NBV), computed using SIENAX (Smith et al., 2001, 2002) which corrects for intracranial size, was used as outcome measure of interest. NBV provides a global marker of brain health and is sensitive to both focal and diffuse atrophy processes in MS.

To condition on disease severity levels, the main feature variable for the normative curves was the baseline Expanded Disability Status Scale (EDSS), which is a discrete non-linear composite measure of disability. Additional covariates included age, sex, disease duration, and clinical subscores relevant to MS disease burden. Image quality metrics (IQMs) were modelled as nuisance covariates and adjusted via counterfactual marginalisation as described in Section 2.2, ensuring that deviation scores reflected biological rather than artefactual variation.

IQMs were derived using the MRIQC pipeline (Esteban et al., 2017) and principal component analysis (PCA) was performed on the full set of 68 IQMs. While the original IQM values were used for model fitting, the first four principal components were retained for counterfactual marginalisation. The choice of four components, together explaining 52% of the total variance, was based on the shape of the scree plot and computational feasibility. Each of the four PCA-derived IQM components was discretised into terciles, resulting in 3^4^ = 81 distinct IQM counterfactual combinations, including the observed IQM profile for each subject. Per counterfactual configuration, we defined a grid point using the median value of each IQM tercile. At each MCMC iteration, the weighted sum of the predicted outcomes across all counterfactual IQM configurations was computed for each subject. Weights were assigned based on the empirical frequency of IQM tercile combinations in the full dataset, ensuring that inference reflects the overall distribution of imaging quality in the data set.

#### Survival model outcome and covariates

The downstream survival module modelled time to first confirmed disability worsening (CDW) as the primary endpoint. A CDW event was defined as an EDSS increase from baseline sustained for ≥ 3months and confirmed at a subsequent visit, using the following thresholds: (i) ≥ 1.5 point increase if baseline EDSS= 0, (ii) ≥ 1.0 point increase if baseline EDSS ≥ 1 and ≤ 5, (iii) ≥ 0.5 point increase if baseline EDSS *>* 5.

Survival covariates comprised clinically relevant demographic and disease characteristics: age, sex, disease duration, relapse count, T1 gadolinium-enhancing lesion count, total T2 lesion volume, timed 25-foot walking test, timed 9-hole peg test, treatment arm, EDSS functional system scores, and MS subtype. IQMs were not re-included here, as their effect was already adjusted for in the normative model.

#### Implementation details

Both the normative BART model and the SoftBART survival model were run with 5-fold cross-validation, 10k burn-in, and 100k posterior draws with 1*/*10 thinning to produce 10k samples for inference. IQM terciles were defined within each fold using the training set only to avoid leakage from the test set. MCMC mixing and convergence were assessed via computation of effective sample size and visual inspection of trace plots, especially of the residual variance parameter.

## 3 Results

### 3.1 Simulations

To benchmark model performance under controlled conditions, we evaluated both the integrated and plug-in approaches using synthetic data with a known generative structure.

Figure 3 compares survival curves across ground truth (dashed), the integrated model (blue), and the CPH model (red). While the integrated model closely recovers the true risk separation, the CPH model underestimates risk especially for earlier times and exhibits confidence intervals that are overly confident.

**Figure 3:**
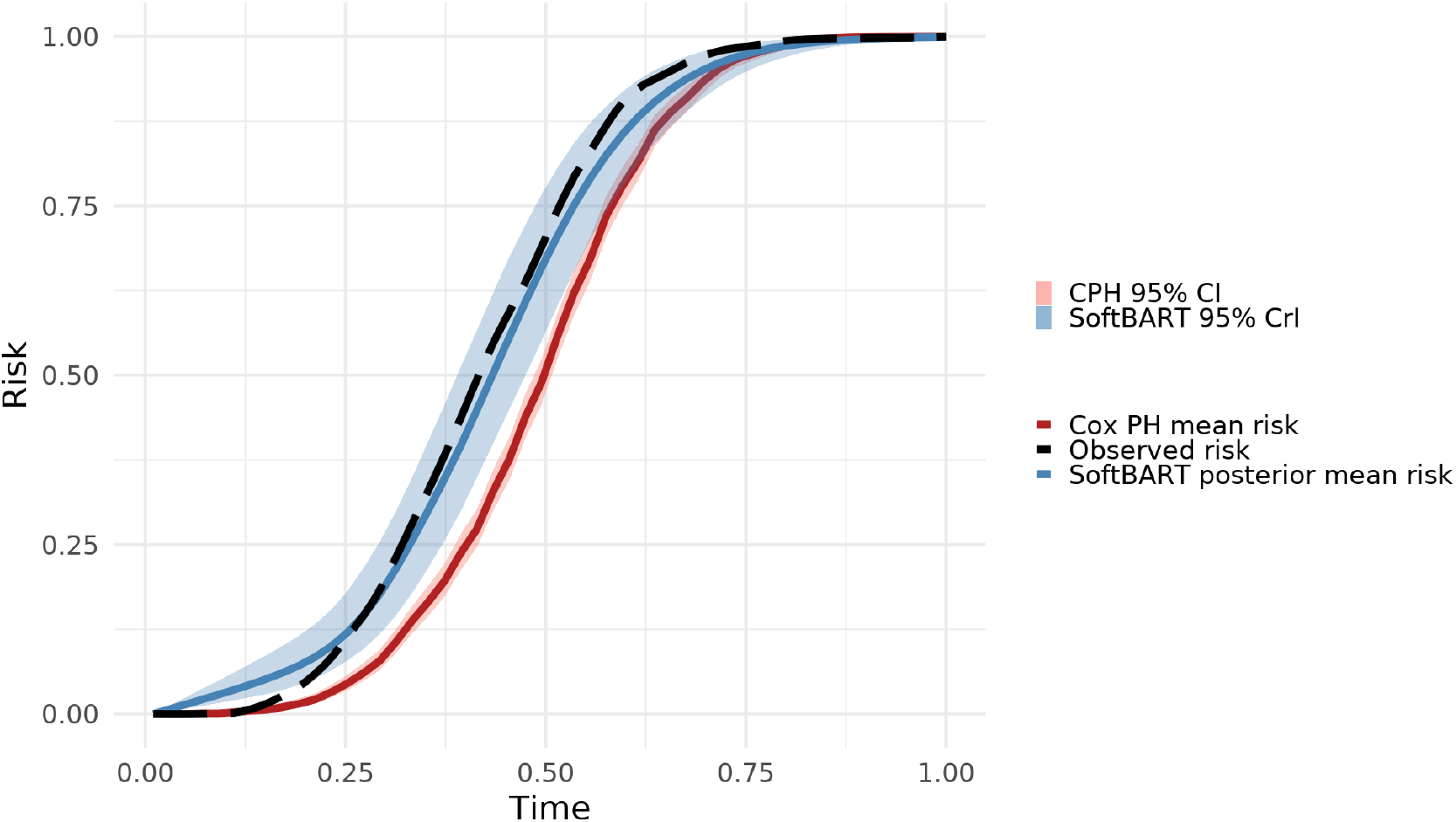
Survival curves with the true risk profile indicated by the black dashed line, the risk profile from our integrated model in blue and the results from a Cox proportional hazards regression in red. Shaded bands indicate 95% credible and confidence intervals, respectively.

#### Quantitative performance metrics

To evaluate our proposed model, we compare it with the CPH model on discrimination and calibration metrics—discrimination refers to the ability of a model to distinguish between outcomes of patients with different risks and calibration assesses the accuracy of prediction (Royston, 2014).

Example calibration plots at one time point are shown in Figure S1. The integrated calibration index (ICI) is 0.109 for the integrated model versus 0.181 for the CPH model. The root mean squared error (RMSE) between true and estimated survival probabilities shows a smaller error of 0.602 for the integrated model versus 0.648 for the CPH model.

Brier scores are lower for the integrated model across all time points (Figure 4, left panel), demonstrating higher prediction accuracy of the integrated model over the CPH model. The right panel of Figure 4 shows the hazard ratio of the two extreme deviation groups (more than one standard deviation from the mean in either direction). The integrated model captures the dynamic risk profile of the ground truth, which the CPH model fails to replicate due to its static formulation.

**Figure 4:**
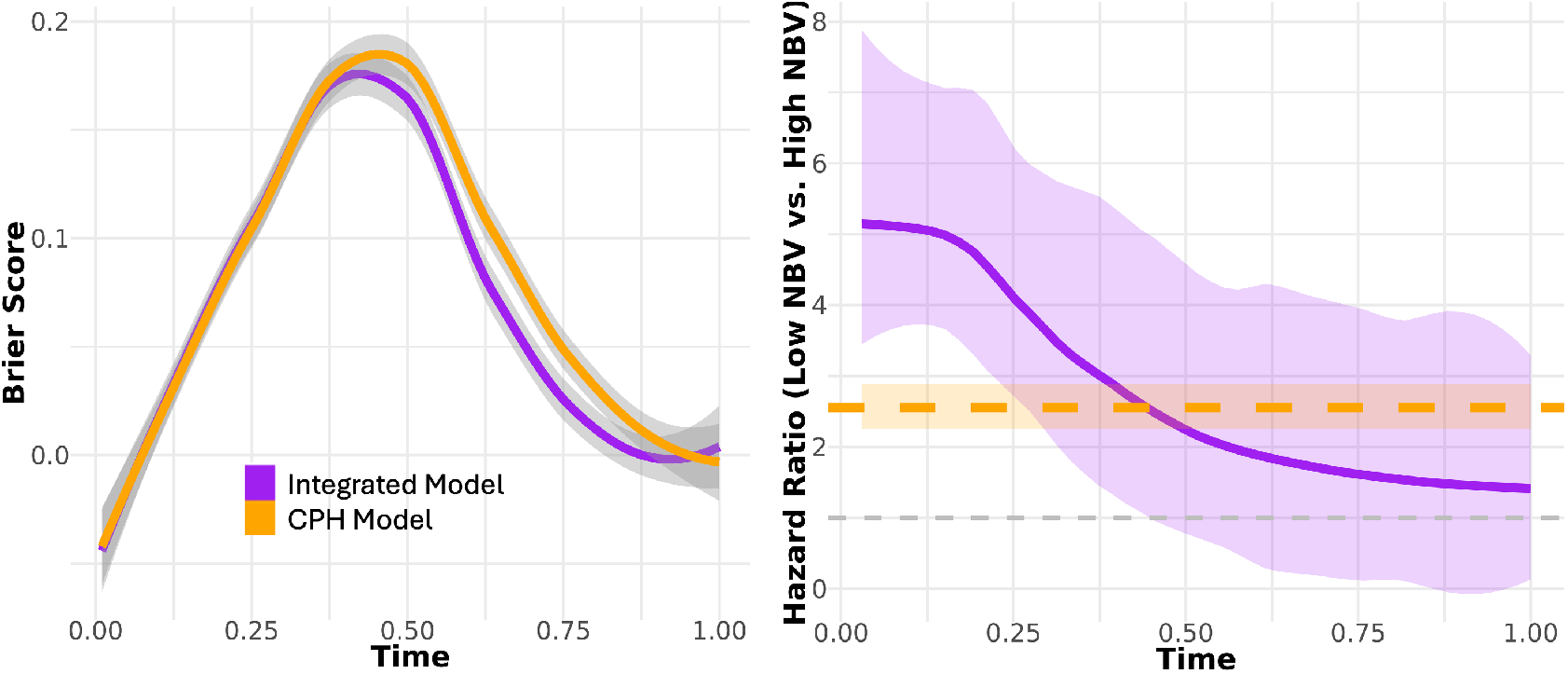
Model evaluations between the integrated model (purple) and the Cox model (orange). The left panel shows Brier scores, with the integrated model performing as well as or better than the CPH model across all time points. The right panel compares hazard ratios for two groups defined as at least 1 standard deviation away from the mean. While the CPH model is limited by the constant hazard assumption, the SoftBART model can accommodate time-varying hazards.

Individual conditional expectation plots and partial dependence plots (Figure S2) illustrate that the integrated model identifies non-linear effects of covariates on survival probability at a selected time point, highlighting the interpretability of SoftBART models in high-dimensional, non-linear settings.

**Table 1:** Summary of quantitative metrics comparing the integrated model and a two-step, plug-in model using CPH regression in the second step.

| Metric | Integrated Model | Two-step Model |
| --- | --- | --- |
| Root Mean Squared Error (RMSE) | 0.602 | 0.648 |
| Integrated Calibration Index (ICI) | 0.109 | 0.181 |
| Brier Score | Lower across all time points | Higher, especially at later times |

### 3.2 Real data

#### Normative patterns of NBV across disease severity

Figure 5 illustrates normative estimates of NBV across the EDSS spectrum. The red solid line depicts the posterior mean trajectory with respect to disease severity. Within each EDSS category, individual normative estimates have been adjusted for potential confounders, including demographic and clinical covariates, and image quality metrics as described in Section 2.2. The population mean is based on an average across subjects with the same EDSS score. Unlike chronological age-based norms, this trajectory isolates disease-related brain volume changes, while accounting for age-related differences in the modelling step. Shaded regions denote posterior predictive uncertainty: green bands encompass *±*1 standard deviation from the conditional mean; the red and blue bands represent lower and upper extremes of deviation scores, respectively, which are more than one standard deviation away from the mean.

**Figure 5:**
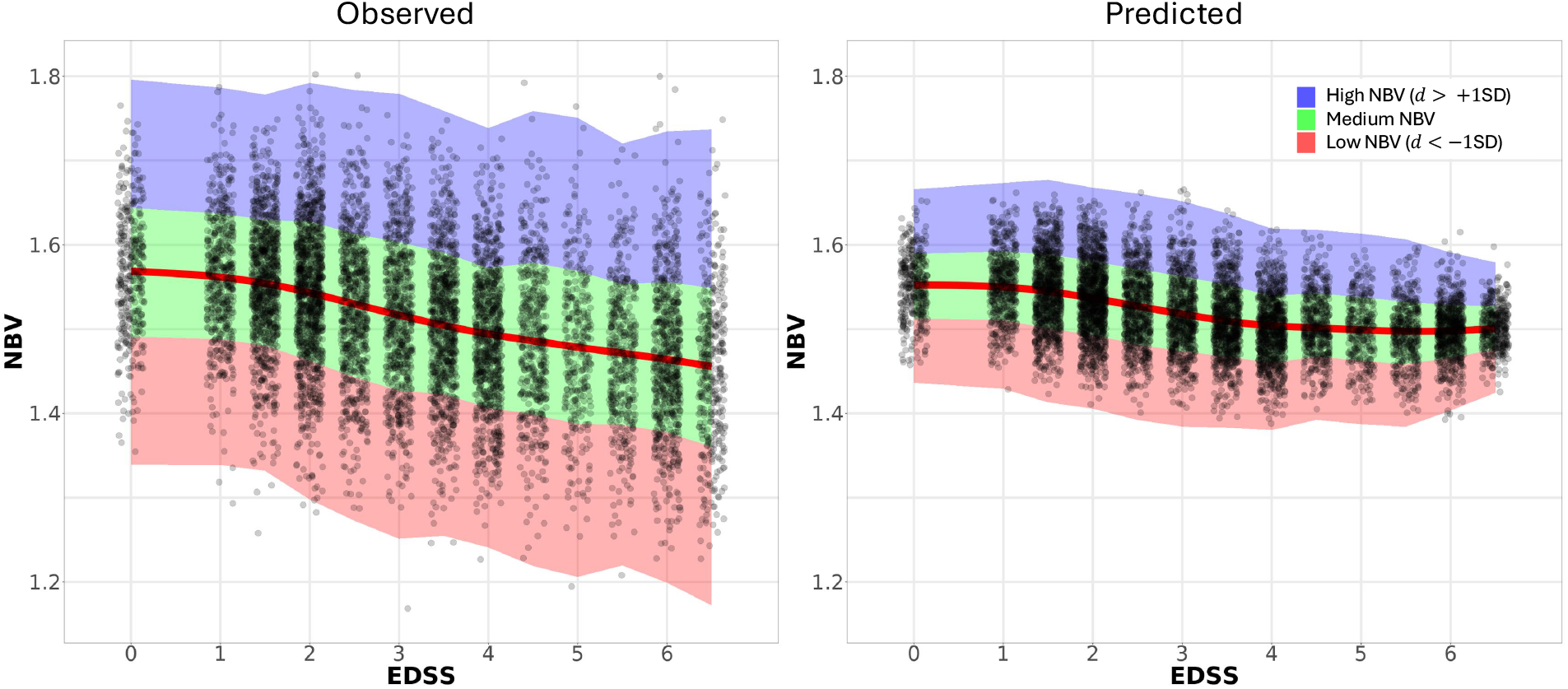
*Left:* Distribution of observed NBV conditional on EDSS level. The red line indicates a smooth fit of the observed population mean. *Right:* Normative curve and individual predicted NBV after marginalisation of nuisance variables. Points in both panels have been jittered around EDSS levels for illustration purposes. The shaded bands indicate regions within (green) or outside (blue and red) one standard deviation of the mean.

Notably, high variability between individuals can be observed across the disease spectrum. Severely affected patients (EDSS 6 or higher) in the highest percentiles of NBV show levels of NBV comparable to individuals close to the population average at EDSS 0. Conversely, individuals in low EDSS categories who fall within the lowest percentiles of NBV may present with lower brain volumes than most MS patients at very high levels of disability.

Figure 5 shows a trajectory that is mostly flat at EDSS 0 to 1.5, consistent with preserved NBV in early MS. A sharper decline is observed at mid-level EDSS scores (2–5), aligning with prior findings that brain atrophy accelerates as MS transitions from predominantly inflammatory to neurodegenerative pathology (Andravizou et al., 2019).

The apparent flattening at higher EDSS levels (*>* 5) may reflect selection bias, as individuals with advanced disability may be under-represented in clinical trials due to physical limitations, cognitive impairment, or difficulty travelling to the scanner site. Note that our data set does not cover late-stage MS (EDSS *>* 6.5).

#### Time-to-event predictions

The traditional two-step approach with a standard plug-in Cox proportional hazards (CPH) model downstream from the normative model resulted in a hazard ratio of 0.564 (0.464 − 0.686, *p <* 10^−8^) when comparing the high NBV to the low NBV group. The modularised model resulted in a hazard ratio of 0.749 (0.683 − 0.824) after the first year and 0.738 (0.669 − 0.818) at day 700, with a median hazard ratio across all time points of 0.751 (0.738 − 0.788); *p <* 10^−6^ at all time points. Reported uncertainty bands are based on a 95% confidence interval for the CPH model and 95% credible intervals for the integrated model.

Figure 6 compares time-to-event predictions from a standard plug-in Cox proportional hazards (CPH) model (left) and the integrated SoftBART model (middle and right panel). Both models included the same set of covariates as described in Section 2.5. The integrated model captures time-varying risk and exhibits greater separation between risk strata defined by deviation scores. In contrast, the CPH model—assuming constant, proportional hazards—underestimates uncertainty and attenuates risk group separation at later time points. Figure 6 also highlights differences between residual-based deviation score estimates (middle panel) and prediction-based scores, with the latter showing better separation between the two extreme deviation groups.

**Figure 6:**
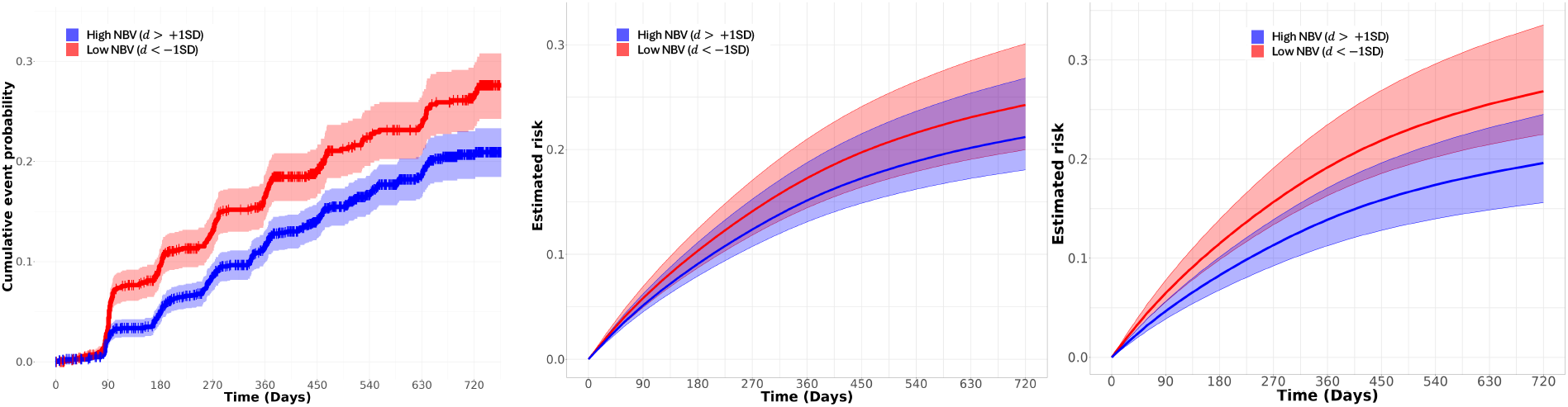
Estimated survival curves using prediction-based deviation scores in a standard Cox proportional hazards model (left) and the integrated model (right). The middle panel shows estimates from the integrated model using residual-based deviation scores instead. The curves show risk profiles stratified by deviation groups (omitting the medium group, i.e. |*d*| ≤ 1SD). Shaded bands indicate 95% uncertainty intervals.

Model evaluation metrics are shown in supplementary Figure S4 and Figure S5. The integrated model consistently achieves lower (better) Brier scores across time, indicating more accurate probabilistic predictions. The individual conditional expectation (ICE) plots in Figure S5 further reveal non-linear covariate effects that are not captured by the linear hazard assumption of the CPH model.

## 4 Discussion

We present a hierarchical Bayesian framework that integrates normative modelling with a second, downstream analysis step via a modularised inference strategy. With a fully joint model computationally out of reach, this modularised approach accounts for uncertainty and allows for unbiased modelling of covariates in the downstream model compared to a two-step plug-in approach.

Leveraging the flexibility of BART and its smooth extension (SoftBART), our approach enables the estimation of robust individual-level deviation scores and for their full posterior distribution to be incorporated into a downstream model. Although our primary focus in this work is on survival analysis, the modular design generalises naturally to alternative downstream tasks such as clustering, classification, and others.

### Normative modelling based on disease severity

Traditional normative modelling has focused on chronological age as the primary reference axis. However, in many clinical contexts disease severity provides a more meaningful stratification. Our approach defines deviation scores relative to normative expectations conditioned on levels of disability. This shift improves interpretability and facilitates clinically relevant characterisations of structural brain health. It aligns with recent work advocating for context-specific baselining in normative inference (Wolfers et al., 2018; Fraza et al., 2024).

Incorporating BART for normative estimation enables flexible modelling of complex, non-linear relationships while accounting for confounding from nuisance covariates. In a real-world application, we apply a principled marginalisation scheme over counterfactual image quality metrics to a large, heterogeneous imaging dataset. This adjustment addresses scanner- and protocol-related variability that would otherwise obscure the biological signal, thereby enhancing the reliability of deviation scores. Crucially, the Bayesian framework allows for principled uncertainty quantification.

### Expectation-based deviation scores

A central innovation of our method is the definition of individual deviation scores as the difference between posterior estimates and the conditional population mean. This contrasts with conventional residual-based scores (e.g., brain age gap), which rely on potentially noisy observed outcomes.

Our approach has several key advantages: First, it avoids conflating measurement error with clinically meaningful variation. Second, it supports principled uncertainty quantification through the posterior predictive distribution. Third, it avoids the mixture of model-based estimates with observed outcomes.

### Modularised inference and uncertainty propagation

A major strength of our framework lies in its use of a cut-posterior inference strategy. This allows posterior uncertainty from the normative module to be propagated to the survival model without feedback. Standard two-step approaches typically ignore the uncertainty in deviation score estimation, leading to biased or overconfident downstream predictions. Our method preserves posterior variability in deviation scores and yields well-calibrated survival estimates.

Additionally, SoftBART enables smooth, time-varying covariate effects, overcoming limitations of piece-wise constant hazard functions in classical Cox models^3^. This is particularly beneficial in clinical trial settings, where interval censoring and structured follow-up schedules can induce temporal artefacts.

We validated our approach on both real-world data from a database of over 8,000 MS patients and under controlled simulations. In both settings, the integrated model outperformed conventional plug-in Cox proportional hazards regression in terms of calibration and prediction accuracy, crucial for any practical application in clinical settings (Riley et al., 2024).

### Limitations and future directions

While powerful, our method is computationally intensive (several hours for the real data application on a single CPU) due to MCMC sampling, particularly in high-dimensional settings and with repeated counterfactual evaluations. GPU-accelerated implementations and variational approximations may help to mitigate this.

By design, the unidirectional flow of information due to the modular framework ignores feedback from the survival model to the normative model. Fully joint models could be explored when such feedback is believed to encode valuable signal, albeit at an even higher computational cost.

Future work could explore generalising this framework from cross-sectional deviation scores to longitudinal modelling of deviation trajectories, i.e. 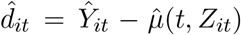, enabling dynamic risk stratification—whether the rate of change in deviation scores is indicative of faster or slower disease progression—and reflecting both within-subject and between-subject abnormality.

Finally, while we focused on survival analysis, the same architecture can support other downstream analyses, such as classification tasks (e.g., cognitive decline prediction) or clustering (e.g., deviation-based disease subtyping), making this a broadly applicable normative modelling framework.

## Conclusion

We presented a novel modular framework that integrates normative modelling with survival analysis using Bayesian tree-based methods. By conditioning deviation scores on clinically relevant covariates and propagating uncertainty through a cut-posterior approach, our method achieves interpretable and robust individual-level risk predictions. This work also extends normative modelling beyond residual-based metrics, offering a powerful tool for modelling disease heterogeneity in a potentially wide range of clinical and neuroimaging settings.

## Data Availability

Code for simulated data is available online at https://github.com/btaschler/NormCutSurv. The clinical data cannot be shared publicly due to ethics and privacy protections.

## Code Availability

Example code for fitting a normative model with BART and a survival model with SoftBART within the modularised framework, together with code for the simulation setup, is available at https://github.com/btaschler/NormCutSurv.

## CRediT Statement

**B.T**.: Conceptualisation, Formal analysis, Methodology, Software, Validation, Visualisation, Writing - original draft, Writing - review & editing; **T.N**.: Writing - review & editing; **H.G**.: Conceptualisation, Methodology, Writing - review & editing.

## Supplementary Materials

### Additional plots for simulation setting

**Figure S1:**
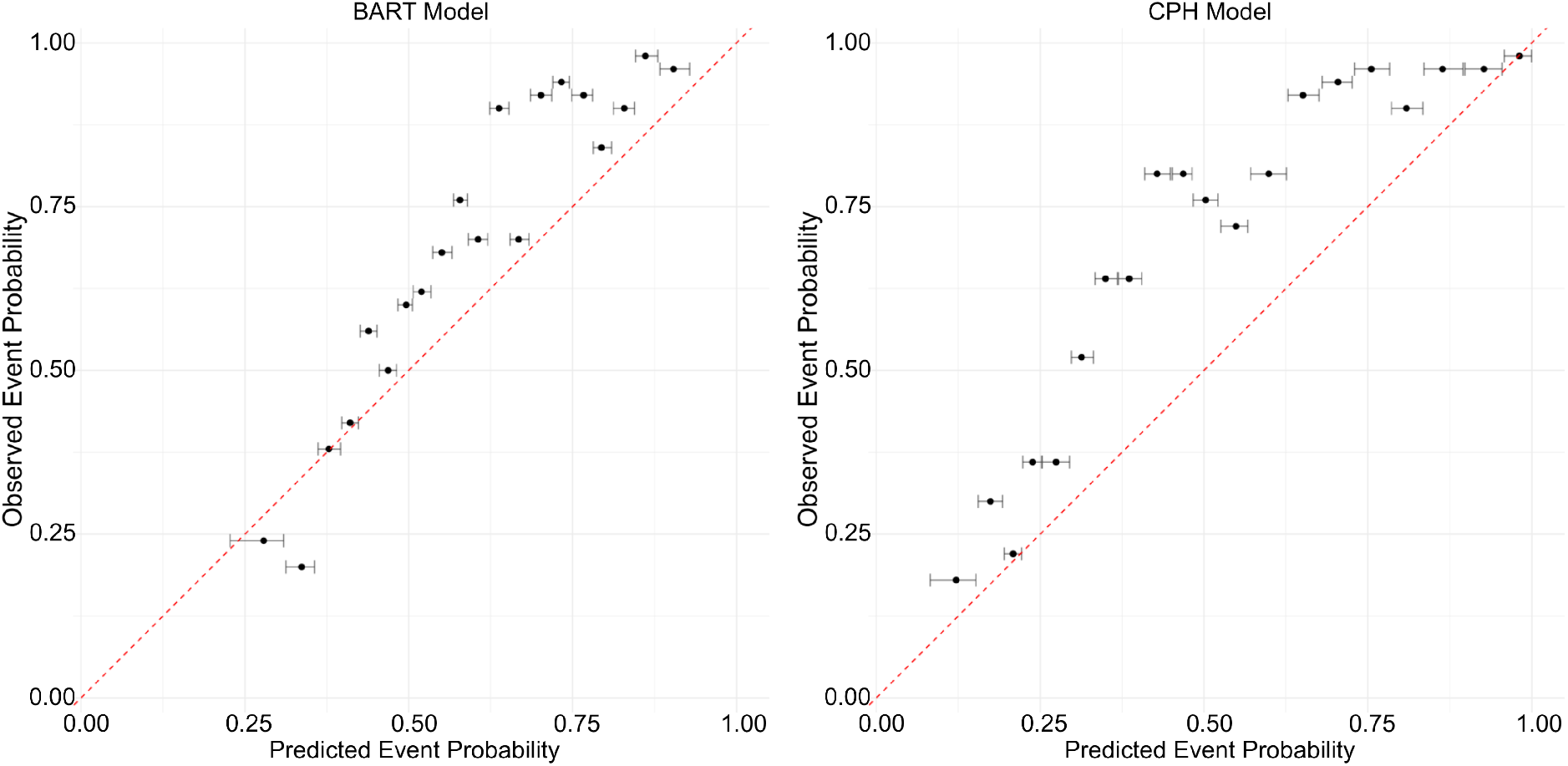
Simulation setting: Example calibration curves at *t* = 0.49.

**Figure S2:**
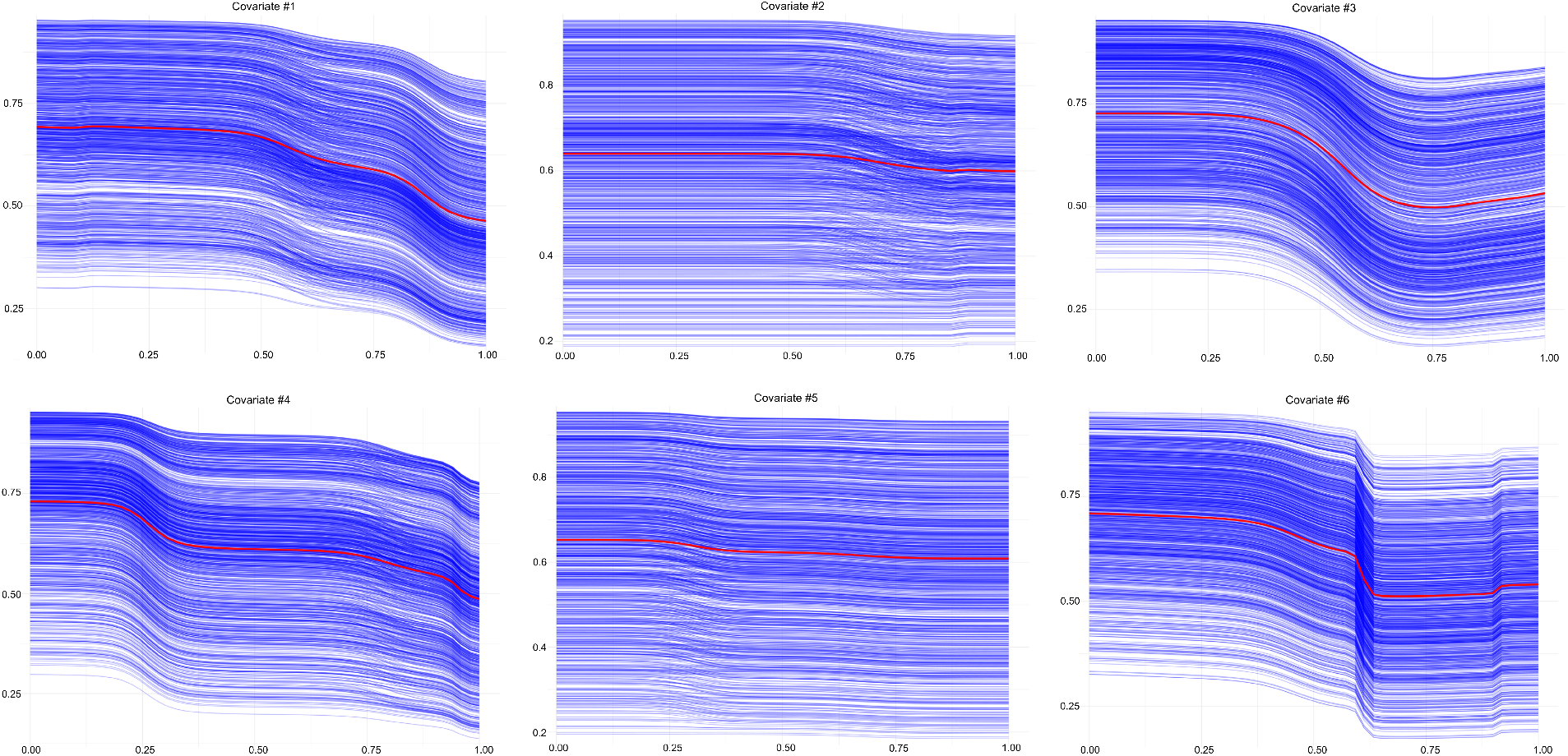
Simulation setting: Individual conditional expectation (blue) and partial dependence plots (red) for all 6 covariates at time *t* = 0.49. The x-axes show standardised values of the covariate, the y-axes show individual event risk.

### Additional plots for real data setting

**Figure S3:**
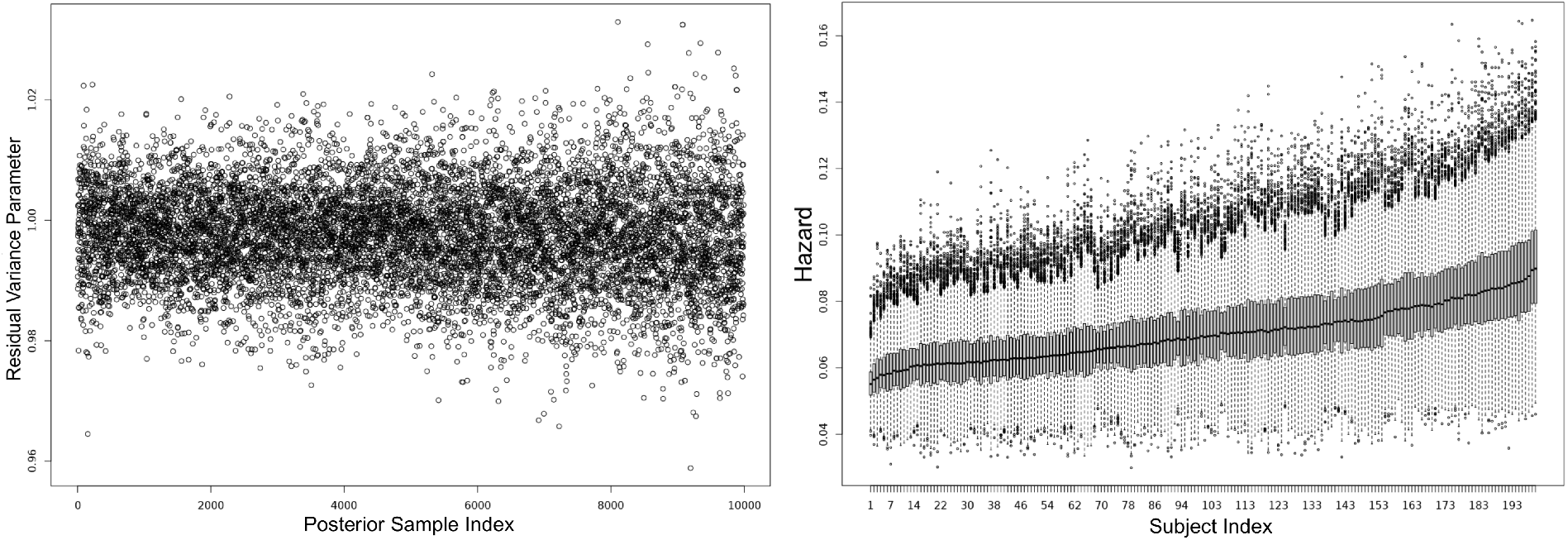
MCMC diagnostics: Trace plot for BART residual variance parameter sigma (left); 200 random subjects and their posterior variance of the predicted hazard.

**Figure S4:**
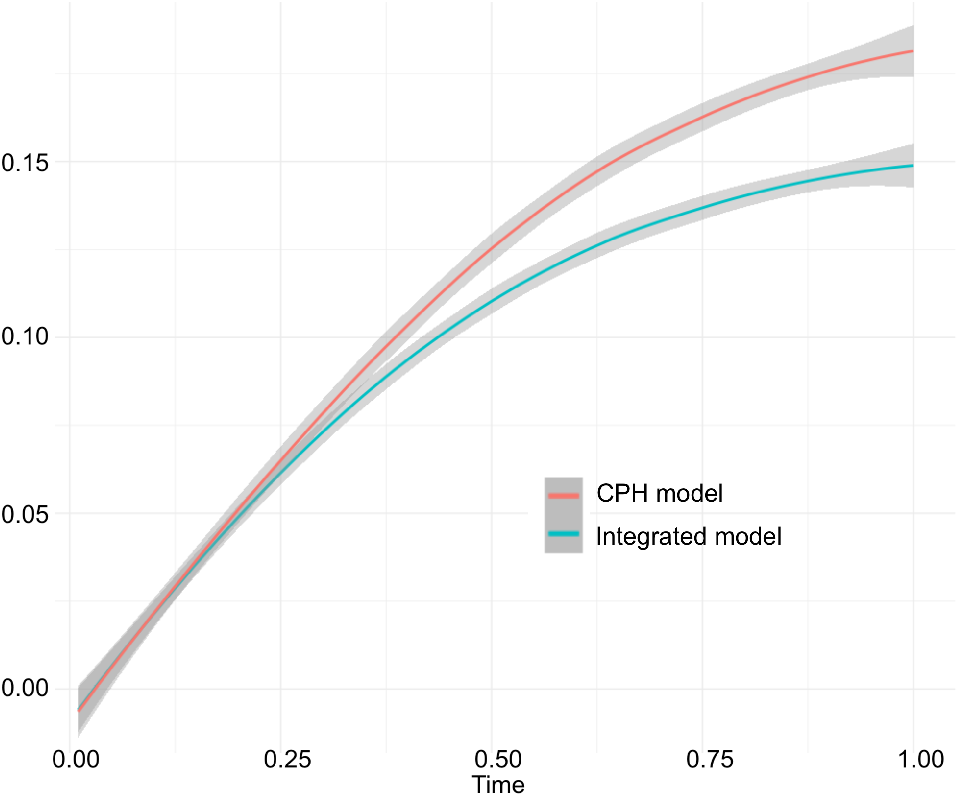
Comparing Brier scores for the standard 2-step approach using a Cox proportional hazards model vs the proposed integrated approach.

**Figure S5:**
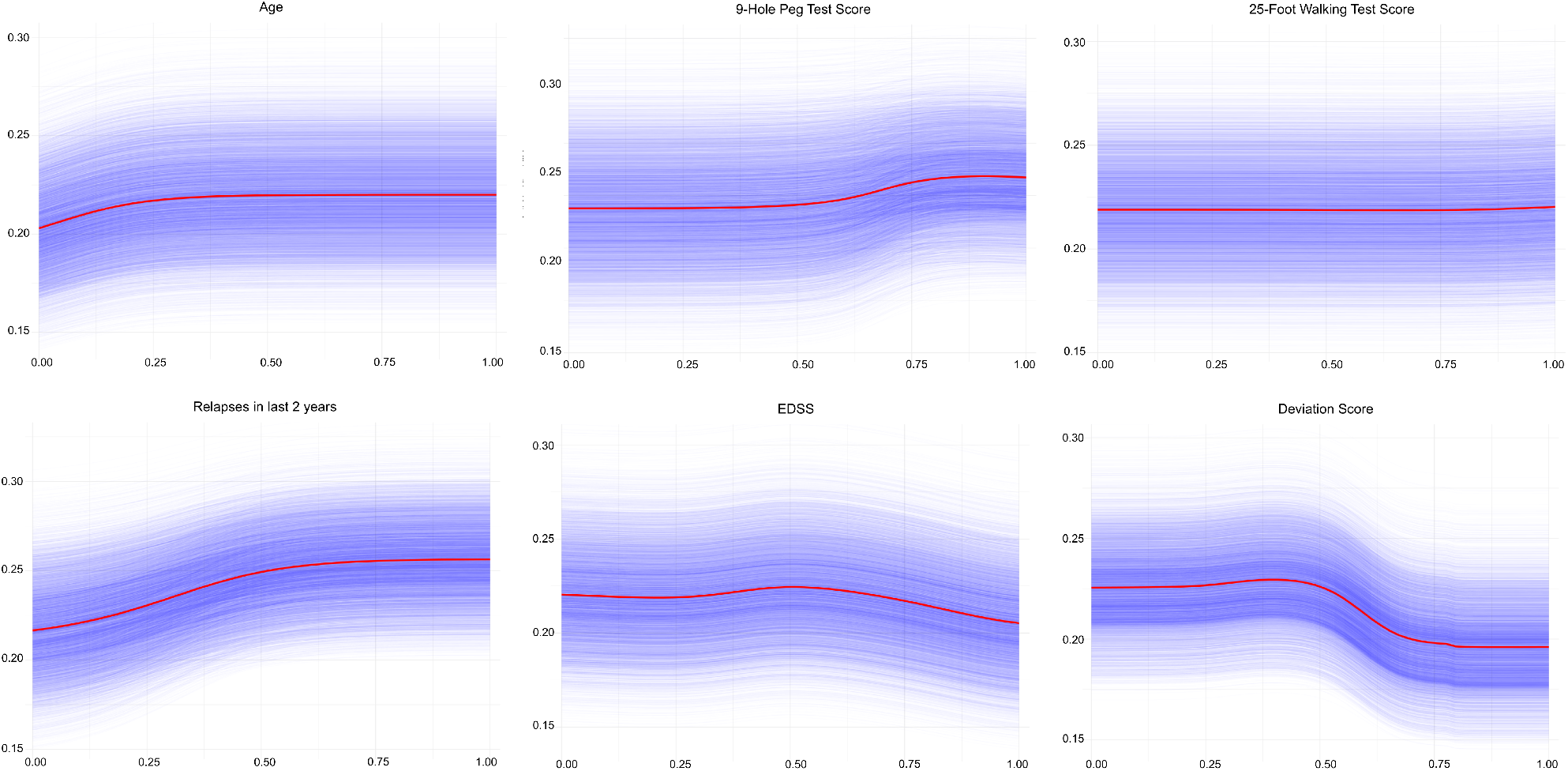
Individual Conditional Expectation (ICE) plots (blue) and Partial Dependence Plots (PDP, red) show how selected covariates influence survival probability at time *t* = 24 months, revealing non-linear effects of covariates. The x-axes show standardised values of the covariate, the y-axes show individual event risk.

## Footnotes

1 In linear settings, marginalisation collapses to the same as conditioning at the mean.

2 Although any variable can in principle take on the role of key feature, for practical purposes we require it to be discrete or discretised with each level sufficiently well represented in the data to allow the estimation of conditional means.

3 Cox regression can incorporate time-varying covariates via start-stop data structures. Each subject’s follow-up is split into intervals where covariates are assumed constant. The partial likelihood is then constructed over these intervals, updating covariate values when they change.

